# Determinants of Collaborative Physician-PA Teams in Ambulatory Care: A Qualitative Study

**DOI:** 10.64898/2026.02.16.26346411

**Authors:** Ashley Nordan, Ian Ward, Melissa Stancil, Gabrielle Schmale, Gayle Bodner

## Abstract

**Background:** The physician assistant (PA) workforce has expanded rapidly in the United States, increasing the importance of effective physician–PA collaboration. Although PAs improve patient outcomes and access to care, the determinants of effective collaboration has not been well studied. North Carolina provides a relevant context due to its growing PA workforce and supervisory regulatory structure, in which physicians retain administrative responsibility for PA supervision across practice settings. This study examines determinants of effective physician–PA collaboration in ambulatory care settings in North Carolina.

**Methods:** Four virtual focus groups were conducted with practicing physicians (n=7) and PAs (n=9) across multiple specialties in NC. Transcripts were analyzed using thematic analysis to identify facilitators and barriers to collaboration.

**Results:** Thematic analysis identified six major themes reflecting relational, organizational, and systemic influences on teamwork. Findings demonstrate collaboration evolves over time through early-career mentorship, continuity of working relationships, and progressive trust development. Differences in professional identity, power dynamics, and misunderstanding of PA scope of practice influenced autonomy and delegation. Systemic factors such as reimbursement structures and organizational supervisory policies hindered efficient teamwork.

**Limitations:** Findings are based on a small, purposive sample within a single state and may not be generalizable to all ambulatory settings or regulatory environments. Perspectives may also reflect self-selection bias among participants with strong views on collaboration.

**Conclusions:** Effective physician-PA collaboration depends on intentional onboarding, role clarity, interprofessional education, and alignment of organizational policies with regulatory standards to support team-based care.

## Introduction

The physician assistant (PA) profession is presently experiencing rapid growth with a 27.8% increase since 2020 and nearly 190,000 certified PAs in the United States healthcare workforce at the end of 2024. [1] In this evolving landscape of healthcare, synergy between physicians and PAs is critical to the delivery of high-quality patient care. Despite documented benefit of PAs regarding patient outcomes and access to care, [2–7] there is currently a lack of research on the specific determinants, qualities and dynamics of truly collaborative physician-PA teams. Better understanding these characteristics could potentially lead to optimizing efficiency and effectiveness of healthcare delivery to meet the growing demand.

North Carolina (NC) is presently the 5^th^ fastest growing state in regard to practicing PAs with more than 11,000 PAs licensed to practice in 2024. [1,8] At the time of this study, physicians in NC have administrative responsibilities for individual PA supervision irrespective of practice characteristics or length of clinical experience. This study focuses on ambulatory care settings to decrease differences in PA role as a variable and because more PAs practice in outpatient settings than inpatient settings. [1]

## Methods

### Research Design

This study used a qualitative study design to examine determinants of collaborative physician-PA teams in ambulatory care settings in NC. As part of a multi-phase research initiative, focus groups with physicians and PAs were conducted to identify key themes related to team dynamics, professional roles and structural influences on collaboration. The study was approved by the institutional review board at Advocate Health - Wake Forest University School of Medicine (Protocol #00122432).

### Population and Sample

Focus group participants were recruited through purposive sampling of existing databases of PA program preceptors affiliated with Campbell University and Wake Forest University. Snowball sampling was also used for intentional recruitment of participants from different specialties, geographic locations and practice sizes.

### Inclusion/Exclusion Criteria

Participants were licensed physicians and PAs practicing in North Carolina for at least one year in the same physician-PA team. Those with less than one year of experience, practicing outside North Carolina, or not working together in the same physical setting at least three days per week were excluded.

### Data Collection and Analysis

Participants were invited by email to complete an eligibility screening based on inclusion and exclusion criteria. Informed consent was obtained electronically through a Qualtrics XM (Provo, UT) survey which also collected demographic information including provider type, years in practice, and specialty.

Semi-structured focus groups were conducted via Zoom (Version 6.6.6) to accommodate participants across different geographic locations. Sessions were audio-recorded, auto-transcribed and reviewed for accuracy, and de-identified prior to analysis. Physicians and PAs were placed in separate focus groups moderated by a non-clinician facilitator to allow participants to speak more freely and eliminate potential barriers to open discussion. Focus group questions were informed by the Interprofessional Education Collaborative (IPEC) competencies and structured to examine interpersonal and systemic factors influencing collaboration. [9]

Two independent coders initially performed inductive and deductive analysis of the four data-rich transcripts, generating a preliminary codebook that included both a priori and in vivo codes with operational definitions. The coders then met to compare coding, resolve discrepancies, and finalize a consensus codebook. All transcripts were subsequently recoded using the consensus version to enhance reliability and ensure consistency across the dataset. All coded data was organized and managed using NVivo 15 software (Lumivero, Denver, CO). Thematic analysis was performed and themes were developed based on code frequency, depth of discussion and interrelationships between codes using the IPEC Core Competencies as the conceptual framework. [9] This process enabled triangulation of findings and refinement of domains.

## Results

### Characteristics of Focus Group Participants

A total of four virtual focus groups were conducted including two PA focus groups (N=9) and two physician focus groups (N=7). The years in practice for participants ranged from 2 to 37 years for PAs (mean = 10.2) and from 3 to 27 years for physicians (mean = 21.1). Participants represented a variety of specialties including family medicine, urgent care, pediatrics, women’s health, internal medicine, emergency medicine, urology, orthopedics and transplant surgery.

### Qualitative Themes

Twenty-four unique codes were categorized as either facilitators or barriers to effective physician– PA collaboration across the four focus groups. From these codes, six overarching themes, including four subthemes, emerged that represented barriers, facilitators, or both, and captured interpersonal and structural factors influencing team collaboration and effectiveness. Although substantial redundancy was observed across groups, full thematic saturation was not reached.

#### Theme #1: Evolving PA Autonomy Through Experience and Trust

Physicians and PAs alike described that early in working relationships, there was often a lack of inherent trust in the training and capabilities of PAs from physicians. Over time, physicians noted increased trust with demonstrated competency on the part of the PA. Likewise, PAs described growth in confidence and independence with less need for a traditional supervisory relationship and desire for a more collaborative relationship with physicians. Lack of continuity with the physician-PA pair negatively impacted the development of this evolving autonomy and trust which included PA turnover and experience working in settings where the PA worked with different physicians (eg. Urgent care or Emergency Department). PAs working with unfamiliar physicians reported repeatedly needing to reestablish their competence, while physicians described the challenges in investing in PAs who later left the practice.

#### Theme #2: Quality of Mentoring Relationships and Onboarding

PAs appreciated early career mentoring and intentional teaching on the part of the supervising physician. Likewise, physicians noted positive experiences working with PAs when a good mentoring relationship was established. Physician participants also perceived themselves as having a responsibility to guide and support PAs in their practice.

#### Theme #3: Differences in Perceptions of PAs

There was a notable difference in how PAs and physicians perceived the PA role, scope, and professional identity. PAs viewed themselves as unique contributors to the healthcare team, emphasizing their distinct training, adaptability, and capacity to enhance patient access and continuity of care. In contrast, physicians often described PAs as extensions of their own practice, focusing on the supervisory relationship.

#### Theme #4: Mutual Respect and Psychological Safety

Mutual respect was repeatedly described as essential to effective physician-PA collaboration by both physicians and PAs. PAs noted that feeling respected and supported by physicians increased their confidence and performance. Likewise, physicians emphasized the importance of maintaining open communication, approachability and availability as well as appreciation for PAs’ experience and receptiveness to feedback to cultivate a strong collaborative relationship. On the other hand, perceived competition between physicians and PAs regarding job security was noted as a barrier to mutual respect between PAs and physicians. Both PA and physician participants reported that this sense of competition was more commonly observed among medical students and residents than among experienced attending physicians.

#### Theme #5: Difficult Power Dynamics

PAs described challenging power dynamics arising when disagreements occurred between PAs and physicians regarding diagnoses or care plans, as well as difficulties implementing care plans with which they personally disagreed. They also reported instances of being caught between two physicians who held differing opinions on patient management, such as conflicts between a supervising physician and a medical director.

#### Theme #6: Systemic and Structural Factors Influencing Collaboration

Both PAs and physicians cited several systemic and structural barriers to collaboration including reimbursement differences for physicians and PAs, organization level policies beyond statutory requirements for supervision, and misunderstandings of PA role and scope.

##### Sub-theme #6a: Differences in Reimbursement

Both PAs and physicians reported reimbursement disparities increased administrative burden for physicians and limited opportunities for true collaboration. This added workload encouraged physicians to be more directly involved in patient encounters primarily for billing purposes rather than clinical need or collaboration. As a result, physicians felt pressure to maintain higher patient volumes while PAs experienced reduced autonomy and efficiency.

##### Sub-theme #6b: Differences in Clinic Structure

Clinic organizational structures and decisions about shared versus separate patient panels was felt to influence the physician-PA collaborative dynamic by both physicians and PAs. Participants highlighted shared panels can promote continuity and teamwork but may also create ambiguity around roles, patient expectations, and follow-up responsibilities. Conversely, separate panels allow PAs to develop independent patient relationships but sometimes reduce opportunities for integrated care. Additionally, distribution of workload was a source of frustration.

##### Sub-theme #6c: Scope Misunderstandings Driving Excessive Oversight

Another theme identified across the focus groups was persistent misunderstandings about PA scope of practice and regulatory requirements, which often led to supervisory structures exceeding what is mandated by law. Both physicians and PAs described how outdated beliefs or institutional policies sometimes imposed unnecessary oversight, blurring the line between what is legally mandated and what is organizationally required. This resulted in perceived increased administrative burden for supervising physicians and limited PA autonomy.

##### Sub-theme #6d: Mixed Perceptions of Mandatory Supervision

PAs and physicians had mixed feelings regarding required supervisory meetings. While PAs felt generally more negatively about the requirement for these meetings, physicians were generally more neutral. While some felt these meetings encouraged collaboration in a good way, others felt that the mandatory nature and required documentation added to administrative workloads. PAs specifically felt mandatory supervision requirements limited collaboration with other, potentially more knowledgeable and experienced, providers outside of their supervising physician.

These themes reflected a wide range of experiences across multiple specialties highlighting variations in PA role, mentoring quality, communication, and the impact of regulatory and organizational requirements on physician-PA collaboration. Exemplar quotes illustrating each theme are included in Table 1. Insights and themes from these qualitative findings directly informed the development of a quantitative survey for NC physicians and PAs, which will be reported in a separate publication.

**Table 1.** Themes, Contributing Codes and Exemplar Quotes.

| Table 1. Themes, Contributing Codes and Exemplar Quotes |  |  |  |
| --- | --- | --- | --- |
| Theme | Facilitator or Barrier | Contributing Codes | Exemplar Quotes |
| 1. Evolving PA autonomy through experience and trust | Facilitator | Evolving PA autonomy<br>Demonstrated competency<br>Teamwork<br>PA turnover | "I think once your physician, can see that you are competent in what you do... you'll start earning the respect [of the physician]." – PA 02<br>"I do feel [that] with experience comes a level of comfort that allows better engagement ... between the physicians and [PAs]." – Physician 07 |
| 2. Quality of mentoring relationships and onboarding | Facilitator | Early career physician mentoring<br>Physician support<br>Physician effectiveness in teaching<br>Open communication<br>Perceived role stewardship | "We should be able to continue to elevate them to our level, regardless... and the [PAs] who don't have [a positive mentoring relationship] are going to find their way somewhere else." – Physician 03<br>"When you come out of [PA] school as a new graduate, you do need someone who's patient and can help you." – PA 05 |
| 3. Differences in perceptions of PAs | Barrier | Role clarity<br>Interprofessional education<br>Perceived role stewardship | "I would have trusted a PA that I taught and trained and mentored much more than some [other PA] at some other practice" – Physician 01<br>"But [physicians] still don't really know exactly what the intent is for a PA, what we're supposed to be able to do, or what the level of training PAs [get] looks like." – PA 08 |
| 4. Mutual respect and psychological safety | Facilitator | Physician Support<br>Mutual respect<br>Open communication<br>Shared values<br>Interpersonal relationships<br>Interprofessional education<br>Perceived competition | "It's the relationship that you make with that person from the moment you become their mentor; the trust and the communication skills that you utilize and build on as you go along." – Physician 03<br>"[The physicians] are never belittling. They never used any terms or [said] between the lines, 'well, you're just a PA'" – PA 07<br>"We treat patients in similar ways. We're kind and we appreciate long-term relationships with patients and their parents. So I think that's the ideal. You believe in the same things, you practice medicine in similar ways." – Physician 01 |
| 5. Difficult power dynamics | Barrier | Practice variability<br>Power dynamics and hierarchy<br>Perceived role stewardship | "If somebody were walking into the practice to see me and they would see my collaborating physician in the hallway, he might just go ahead and take them and see him himself." – PA 06<br>"I remember a few times when I'd feel very different about the treatment plan of a patient versus my supervising physician, and ultimately, he left it up to me. But you almost feel like you're betraying someone because they helped mold you into who you are." – PA 05<br>"I've kind of gotten in between two doctors, having two different ideas of how you should treat a patient... then you're like, 'Well, I don't know which to do' and it can be really uncomfortable and awkward." – PA 03 |
| 6. Systemic and structural factors |  |  |  |

Determinants of Collaborative Physician-PA Teams in Ambulatory Care: A Qualitative Study
| influencing collaboration |  |  |  |
| --- | --- | --- | --- |
| 6a. Reimbursement disparities | Barrier | Billing<br>Competing priorities | “Our [physician] used to say, when we first started, that he is required to see all Medicare patients first before we do... it made it hard to establish new patients..” – PA 06<br>“Some of the commercial insurance plans only reimburse us 85% when they're seen by a PA...So that's a problem in my mind, right? They're doing the same work that a physician would be doing.” – Physician 04 |
| 6b. Differences in clinic structure | Both | Teamwork<br>Shared values<br>Competing priorities | “The second PA I worked with... was probably my favorite of my work interactions because they were primarily assigned to me, and we functioned like a team to take care of my panel of patients.” – Physician 05<br>“It's challenging when a patient sees different providers who manage things differently.” – PA 09 |
| 6c. Scope misunderstandings driving excessive oversight | Barrier | Misunderstandings of PA scope and regulation | “I don't know if that ever was really a rule, that you had to sign all the notes, but that's just what I've always done, because it's what I was told I was supposed to do.” – Physician 02<br>“If you have a corporate structure that either, doesn't know how to treat [the PA] or doesn't view [the PA] as making the appropriate decisions that we're able to make, I think that can contribute to either poor or less than ideal outcomes as well.” – PA 09 |
| 6d. Mixed perceptions of mandatory supervision | Both | Required supervisory documentation of meetings<br>PA practicing under the physician's license | “Sometimes I feel like [required supervisory meetings] are just checklist things... because... we collaborate all the time” – Physician 07<br>“[The PA and I] schedule quarterly meetings where we debrief, and we've tied in some of the formal documentation that's required... It's gone well and it aids in the interaction.” – Physician 05<br>“I think that we speak to our supervising physicians so much that the fact of a [required supervisory] meeting is just time that we could be doing other things.” - PA 09 |

## Discussion

This qualitative study contributes to the limited literature available describing physician–PA collaboration as a developmental process rather than a fixed supervisory model. Participants’ descriptions of progression from early-career oversight to mature collaborative partnerships align with prior work on PA transition-to-practice and onboarding frameworks, which emphasize the importance of structured mentorship, role clarity, and graduated autonomy. [10–13] The consistency with which both PAs and physicians identified a point at which rigid supervision became unnecessary suggests that collaborative practice is best interpreted as an evolving professional relationship shaped by experience, trust, and continuity rather than strict regulatory standards alone.

A notable finding was the role of trust formation in shaping collaboration. Reports of physicians extending less inherent trust to PAs than to physician colleagues, which required trust to be earned through demonstration of competence, helps explain why early career mentoring emerged as such a critical facilitator. Mentorship served not only as a means for clinical support but also as a mechanism for bidirectional trust building. This enabled physicians to reframe expectations of PAs and PAs to gain confidence and autonomy. Where there was a lack of continuity, this process repeatedly reset which reinforced supervisory models and slowed progression toward true collaborative practice. These findings align with literature on interprofessional feedback processes which suggests that structured, ongoing, bidirectional peer feedback can enhance role clarity, communication and team performance. [11] By reinforcing shared observations and constructive evaluation early in practice, peer feedback may serve as a means by which to accelerate trust building and support ongoing collaboration between physicians and PAs.

Differences in conceptualization of the PA role further shaped these dynamics. PAs viewed themselves as independent contributors who enhanced access and continuity of care which contrasted with physicians’ tendency to view PAs as extensions of their own practice. This difference reflects longstanding debate in the literature regarding the historical labeling of PAs as “physician extenders” which may continue to influence role expectations among more experienced clinicians. [15] When combined with organizational policies that limit PAs’ ability to practice at the top of their license, this misalignment appeared to drive inconsistent delegation patterns and unnecessary oversight which consequently increased physician administrative burden while negatively impacting PA efficiency.

At the interpersonal level, findings strongly reinforce existing interprofessional literature emphasizing mutual respect, psychological safety, and open communication as core elements of effective team-based care. [9] These relational facilitators closely mirror the competencies outlined by the Interprofessional Education Collaborative, particularly in domains of values and ethics, roles and responsibilities, and interprofessional communication. At the same time, participant accounts of perceived competition, conflicting directives, and uneven workload distribution illustrate how hierarchical structures can undermine these competencies when not effectively addressed. Notably, both PA and physician participants emphasized this perceived competition was most commonly observed among medical students and residents as opposed to more experienced attending physicians. This further suggests that early professional socialization and limited exposure to PAs during training may contribute to misunderstandings or lack of understanding about the PA role on the healthcare team. [16–18]

Systemic factors at the structural and policy level also emerged as highly influential to the success of physician-PA collaboration. Reimbursement models, organizational supervision requirements exceeding statutory standards, and clinic structures that did not align with professional roles functioned as systemic constraints and barriers to collaboration. These findings suggest that collaborative practice is not solely a function of interpersonal or professional intent, but is significantly molded by institutional and policy contexts which can either facilitate or inhibit team dynamics.

## Limitations

This study has several limitations that should be acknowledged. First, although both physicians and PAs contributed valuable insights, there was a significant difference in mean years in practice between the two groups. This may have influenced perspectives on PA autonomy, supervision, and collaboration. Despite this difference, the overall range of practice experience was similar providing representation across career stages for both professions.

Generalizability of findings is limited by the composition and context of the sample. Physicians were predominantly practicing in primary care, whereas the PAs represented a broader mix of specialties creating an imbalance in practice area representation that may limit comparability between groups. In addition, the study reflects experiences from North Carolina where PA practice laws and professional norms may differ from those in other states.

Another limitation is that full thematic saturation was not achieved. Although substantial redundancy emerged across the first three focus groups, the final group generated one additional code suggesting that additional data collection may have revealed further nuances in perspectives on collaboration. Additionally, the findings reflect participant perceptions rather than objective measures of collaboration, clinical effectiveness or patient outcomes. Focus group dynamics may have also influenced participants’ willingness to share opposing or sensitive perspectives.

Lastly, the sample consisted of a purposive group of PAs and physicians who mostly serve as preceptors or educators. Given this background, these participants, especially the physician participants, may be more supportive of PAs or more familiar with interprofessional collaborative practice than non-preceptor clinicians. This may have resulted in responses that are more positive toward the PA role than those found in the general clinician population.

### Future Research

Future research evaluating structured onboarding programs or physician training focused on supervision and interprofessional collaboration could provide evidence-based models for implementation. Comparative studies across states with varying regulatory requirements may help identify policies that most strongly support effective collaboration. Quantitative measurements in a larger study population could also validate findings and expand on themes identified in this study.

## Conclusion

In conclusion, this study demonstrates effective physician-PA collaboration in North Carolina develops through a pathway of early career mentoring, continuity, progressive autonomy and trust-building, which are all influenced by organizational and regulatory contexts. Although grounded in North Carolina’s practice and supervisory environment, these findings reflect common dynamics present in other states and health systems that employ PAs within team-based care models.

These findings suggest several actionable strategies for health systems. First, standardized onboarding and early career mentorship protocols may help establish the foundational trust and competence needed for efficient collaboration. Second, prioritizing continuity in team assignments so that physicians and PAs work together consistently over time can facilitate the development of shared workflows and mutual confidence. Third, inclusion of PA role education into physician training and orientation may reduce misconceptions about PA capabilities and better support shared decision making. Implementing these strategies may enhance team-based care, expand access, and strengthen workforce sustainability in ambulatory care settings.

## Data Availability

All data produced in the present study are available upon reasonable request to the authors

## Acknowledgements

A pre-press copy of this manuscript has been posted to the medRxiv.org pre-print server. This manuscript was created as part of graduation requirements for the DMSc program of Wake Forest University School of Medicine Physician Assistant Studies. The authors have no relevant conflicts of interest.

## Notes

### Competing Interest Statement

The authors have declared no competing interest.

### Funding Statement

The study was partially funded by an internal faculty research grant from Campbell University College of Pharmacy & Health Sciences.

### Author Declarations

IRB of Advocate Health - Wake Forest University School of Medicine gave ethical approval of this work.

